# Theory of Mind and Moral Decision-Making in the Context of Autism Spectrum Disorder

**DOI:** 10.1101/2021.03.13.21253459

**Authors:** Jessica Ellen Ringshaw, Katie Hamilton, Susan Malcolm-Smith

## Abstract

Social impairment in Autism Spectrum Disorder (ASD) has been linked to Theory of Mind (ToM) deficits. However, little research has investigated the relationship between ToM and moral decision-making in children with ASD. This study compared moral decision-making and ToM between aggregate-matched ASD and neurotypical boys (*n=*38 per group; aged 6-12). In a third-party resource allocation task manipulating recipient merit, wealth and health, neurotypical children allocated significantly more resources to the morally deserving recipient, suggesting equitable allocation. A comparatively larger portion of the ASD group allocated equally. ToM emerged as a predictor of moral decision-making. We suggest that ToM (cognitive empathy) deficits may underly atypical moral decision-making in ASD by limiting the integration of empathic arousal (affective empathy) with moral information.

## Theory of Mind and Moral Decision-Making in the Context of Autism Spectrum Disorder

Autism Spectrum Disorder (ASD) encompasses a wide range of clinical presentations with symptoms that are expressed along a continuous spectrum of severity (APA, 2013; Lauritsen, 2013). Although an ASD diagnosis is based on two domains of impairment, it is primarily regarded as a disorder of social impairment (APA, 2013; Kim & Lord, 2013). Diminished social competency in ASD has consistently been linked to Theory of Mind (ToM) deficits (Baron-Cohen, 1995; Baron-Cohen et al., 1985; Mazza et al., 2017)

### ToM in ASD

Theory of Mind (ToM) is broadly defined as the ability to attribute mental states such as beliefs, desires, perspectives and intentions to others, and to recognise that these are independent of one’s own mental states (Baron-Cohen, 1995; Baron-Cohen et al., 1985). In neurotypical individuals, ToM and its precursors start emerging as young as 14 months and tend to follow a predictable developmental trajectory throughout childhood (Low & Perner, 2012; Wellman et al., 2001; Wellman & Liu, 2004).

In contrast, ToM deficits have consistently been associated with ASD and become noticeable in behaviour from an early age (Baron-Cohen, 1995; Baron-Cohen et al., 1985; Happé, 1993; Happé & Frith, 1996). Many children with ASD fail formalised ToM tests (Baron-Cohen et al., 1985; Mazza et al., 2017), and it has been proposed that ToM development may be delayed or even deficient in ASD (Hoogenhout & Malcolm-Smith, 2014). The direct relationship between ToM and social competence in ASD (Aljunied & Frederickson, 2011; Charman et al., 2000; Fombonne et al., 1994; Lerner et al., 2011; Travis et al., 2001) remains relevant in understanding the social difficulties experienced by this group.

### Theory of Mind and Moral Decision-Making

Moral decision-making falls under the umbrella of social decision-making due to the social environment within which moral decisions are made, and the reliance of this process on the evaluation of socially defined values (Greene & Haidt, 2002; Rilling & Sanfey, 2011). It is defined as the process by which one makes choices based on moral judgements (Reniers et al., 2012). In turn, moral judgement involves assessing behaviours that revolve around socially constructed norms and moral principles (Ciaramelli et al., 2007; Prehn et al., 2008).

### Developmental Trajectory of Moral Decision-Making

The development of moral decision-making in neurotypical samples has been characterised by changes in sharing behaviour with age as children start incorporating social norms into their perception of fairness. At the age of 15 months, children demonstrate an expectation for fairness (Schmidt & Sommerville, 2011) and by the age of 3, they express a desire for equal distributions in third-party resource allocation tasks (Baumard et al., 2012; Smith et al., 2013). By the age of 6, they begin making judgements about whether recipients are deserving with regard to merit and well-being (Damon, 1977; Rizzo et al., 2016). Furthermore, slightly older children (aged 7 to 8) rectify inequality in their judgements and allocations, demonstrating a preference for equitable distributions over equal distributions in scenarios represented by inequality (Kienbaum & Wilkening, 2009; Malti et al., 2016; Rizzo & Killen, 2016).

While moral decision-making behaviour is characterised by significant change over the course of development, the underlying mechanism for this is less understood (Cowell et al., 2015). The cognitive capacities suggested to facilitate sharing include ToM (Takagishi et al., 2010) and executive functions (Liu et al., 2016; Smith et al., 2013). However, there is limited evidence for a clear relationship between generosity and the latter (Aguilar-Pardo et al., 2013; Smith et al., 2013). Therefore, from a developmental point of view, ToM is a more likely candidate underlying age-related changes in moral decision-making.

### The role of empathy in moral decision-making

Research suggests that ToM is implicated in most (but not all) moral judgements due to its involvement in higher-order moral emotions (Greene & Haidt, 2002; Greene et al., 2004; Moll et al., 2002). In examining the pattern of overlapping emotion and cognitive neural circuitry in moral decision-making, it is clear that integrative empathy pathways are involved (Decety et al., 2012). According to Decety (2011), “empathy” encompasses affective empathy, cognitive empathy and self-regulation. While affective empathy refers to emotional sharing or arousal, cognitive empathy overlaps with ToM and refers to the ability to infer the mental states of others for improved emotion awareness and understanding. Self-regulation enables the control of emotion, affect and drive (Decety, 2011). In contrast to affective empathy which is present from infancy, cognitive empathy and self-regulation develop with age (Decety & Jackson, 2004; Decety & Meyer, 2008).

This model proposes that empathic concern is enabled by appropriately regulated affective empathy and the mediatory role of cognitive empathy (ToM; Decety, 2011). In considering this 3-pillar model of empathy within the context of moral cognition, empathic concern is coined as motivational empathy and refers to the urge to help or care. It is suggested that affective perspective-taking (the integrated effect of cognitive and affective empathy) elicits empathic concern and, potentially, prosocial behaviour (Batson, 2012; Decety & Cowell, 2014a, 2014b; Van Lange, 2008)

While this serves as a basis for understanding the role of empathy (including cognitive empathy) in morality, it is an area of ongoing research. Despite neuroimaging evidence for a relationship between ToM and moral decision-making (Decety & Cowell, 2014a, 2014b; Decety et al., 2012; Young et al., 2007; Young & Dungan, 2012), behavioural studies are necessary to ascertain the phenotypic presentation. Thus far, most behavioural studies have focused on abstract moral reasoning using convoluted tasks that lack external validity (Kahane et al., 2015). However, with the development of simple and scientifically interpretable tasks, complicated and contradictory results have emerged (Cowell et al., 2015; Takagishi et al., 2010). Further studies using similar tasks are required to validate the role of ToM in moral decision-making and to fully understand its mechanism. This is particularly relevant to ASD research given that its presentation often includes ToM deficits.

### Moral Decision-Making and ASD

Given the well-known ToM deficits associated with ASD and the pathognomonic social impairment central to the disorder, some studies have suggested that moral decision-making may be atypical in this clinical group. This is supported by evidence of atypical patterns of neural activation (Schneider et al., 2013) as well as concrete moral reasoning and inflexible rule-based condemnations (Shulman et al., 2012).

Furthermore, some studies have linked atypical moral judgement and inappropriate moral justifications with ToM deficits in high-functioning adults with ASD. For example, adults with ASD and ToM deficits had greater difficulty distinguishing between accidental and intentional harm (Moran et al., 2011), demonstrated diminished capability in rating injury severity subsequent to making blame judgements in unfamiliar social scenarios (Salvano-Pardieu et al., 2016), and poorly evaluated the seriousness of moral transgressions (Zalla et al., 2011). ToM deficits in adults with ASD have also been linked with an increased likelihood of making utilitarian judgements in personal dilemmas and reporting reduced distress in perceiving such scenarios (Gleichgerrcht et al., 2013). Additionally, due to ToM deficits, adolescents with ASD tend to make blame judgements based on the physical consequences of action rather than the intention of the perpetrator (Salvano-Pardieu et al., 2016), and have difficulty inferring willingness to forgive (Rogé & Mullet, 2011).

However, these findings are not consistent throughout the literature as there is evidence for contrary results (Blair & James, 1996; Leslie et al., 2006; Patil et al., 2016; Sally & Hill, 2006). For example, new and advanced research measuring both explicit (verbal) and implicit (eye-tracking) responses has revealed that children with ASD are in fact able to understand a perpetrator’s intention in making third-party judgements (Li et al., 2019). However, they demonstrated milder (less negative) explicit judgements and had an atypical response pattern characterised by increased sensitivity to damaged objects rather than hurt people. This indicates that an overriding interest in non-social stimuli may be more relevant than deficits in basic ToM skills (i.e. intention detection). However, it does not exclude the possibility of higher level ToM skills being necessary for moral decision-making by driving an intricate mechanism that integrates affective empathy with social information (Li et al., 2019). Overall, in comparison to neurotypical research, there is limited reliable evidence from which to draw conclusions regarding the role of ToM in morality in ASD, particularly within the paediatric population.

### A Potential Theoretical Framework for Predicting Moral Decision-Making in ASD

Despite the lack of research on morality in ASD, some theories attempt to explain how social decisions tend to made by this group. The Empathizing-Systemizing (E-S) Theory proposes that some individuals have hyper-developed “empathising brains” (Type E) while others have hyper-developed “systematising brains” (Type S), and a small proportion have “balanced brains” (Type B) (Baron-Cohen et al., 2003). The E-S Theory claims to explain sex differences whereby, on average, more males have Type S brains while more females have Type E brains. In turn, the Extreme Male Brain (EMB) Theory of Autism suggests that the male pattern is exaggerated in ASD (Baron-Cohen, 2002).

“Systemizing” refers to the process of predicting and controlling phenomena that are lawful and deterministic (Baron-Cohen, 2009; Baron-Cohen et al., 2003). Given that people are not rule-governed, empathising is more effective in predicting human behaviour. This requires perspective-taking (ToM) and an understanding that the relationship between a person’s mental state and behaviour is not lawful. The E-S and EMB theories emphasize that ASD is associated with a lack of “empathy” and an increased tendency to systemize. Given that empathy is a multidimensional construct (Dziobek et al., 2008; Fan et al., 2014), it is proposed that ASD is associated with an inability to understand and predict mental states (ToM deficits; cognitive empathy) rather than a general lack of empathic arousal or concern (Fan et al., 2014). To compensate, individuals with ASD may resort to predicting the behaviour of systems rather than people when faced with the social world (Baron-Cohen et al., 2005). In line with this hypothesis, Kennet (2002) suggested that ASD individuals are likely to adopt a Kantian moral approach by applying rules without much regard for the agent’s emotion or the ultimate good of the action.

### Rationale

In neurotypical individuals, the development of ToM and moral decision-making is relatively well understood. Furthermore, ToM has been proposed as an important cognitive component of moral decision-making. Given the documented ToM deficits in ASD, it is likely that moral decision-making may be atypical. However, very little evidence-based research has explored the relationship between ToM and moral decision-making in ASD, particularly within the paediatric population. The relevance of such research lies in its ability to provide insight into the mechanisms underlying aspects of social impairment in ASD. The aim of this study was to compare moral decision-making in ASD and neurotypical children using a third-party resource allocation task, and to determine the role of ToM in this. We hypothesized (hypothesis 1) that neurotypical children would use the social information provided and, therefore, allocate significantly more resources to the morally deserving recipient (equitable allocation) across conditions of inequality. In contrast, we expected children with ASD to allocate resources more equally based on the likely systematic use of mathematical and moral rules pertaining to fairness. We also hypothesized (hypothesis 2) that ToM would be a significant predictor of moral decision-making for all children.

## Method

### Design

A cross-sectional, relational design was used to compare moral decision-making patterns in ASD and neurotypical children. ToM was examined as a potential predictor of moral decision-making in both groups. Children with ASD were recruited from the UCT Autism Research Group’s database of families willing to participate in research, as well as several ASD-specific and special-needs primary schools in the Western Cape. These participants comprised the clinical group. Neurotypical children for the control group were recruited from public primary schools in Cape Town. We recruited all children from English-medium schools and parents were required to be fluent in English. Data collection took place in quiet classrooms or, in select cases, we assessed participants in a quiet room at their homes.

### Participants

A total sample of 76 boys between the ages of 6 and 12 were included in the study, with 38 participants per group. Aggregate matching was conducted across groups based on demographic information (age and socio-economic status(SES)). Participants in both groups were from schools serving low to middle income communities. All participants in the ASD group had an existing ASD diagnosis which was confirmed by an *ADOS2 (Autism Diagnostic Observation Schedule, Second Edition;* Lord, 2012) assessment prior to further testing. Non-verbal ASD children were excluded given that ToM can only be extensively assessed in verbal children (Hamilton et al., 2016). Exclusion criteria for the ASD group included a history of head injury or diagnosed neurological disorder such as epilepsy. However, Attention-Deficit and Hyperactivity Disorder (ADHD) and anxiety disorders did not constitute exclusion due to the high co-morbidity with ASD (Peacock et al., 2012; Simonoff et al., 2008). Exclusion criteria for the neurotypical group included diagnosed neurodevelopmental disorders, neurological disorders and a history of infantile meningitis or head injury.

### Measures

#### Demographic survey

Prior to testing, the parents/legal guardians of all participants completed a demographic survey. This provided data regarding child age and SES (indicated by the annual Total Family Income). The demographic survey data was used to aggregate match ASD and neurotypical groups based on age and SES. Additionally, demographic variables (i.e. age and SES) were included in analyses to account for their potentially predictive effect on moral decision-making.

#### ASD diagnosis

All children who were recruited into the ASD group were assessed using the *Autism Diagnostic Observation Schedule*, Second Edition (*ADOS2*; Lord, 2012) in order to validate and confirm an ASD diagnosis. The *ADOS2* is a semi-structured, standardised instrument that is considered the gold standard for the observational assessment of ASD in clinical diagnostics and research. The verbal modules of the *ADOS2* were administered by a certified research-reliable *ADOS* administrator on the team, and the assessment produced a score representing overall ASD presence.

#### Verbal comprehension

To ensure comprehension difficulties did not confound the ToM results, we screened for this prior to the administration of the *UCT ToM Battery* with the *Comprehension of Instructions* task from the *Developmental Neuropsychological Assessment*, Second Edition (NEPSY-II; Brooks et al., 2009). This was used to assess whether children could follow two-stage commands that involve the comprehension, processing and execution of instructions. The *NEPSY-II* has been translated into several languages and both editions have acceptable psychometric properties based on samples from Western and non-Western countries (Abedi et al., 2012; Mulenga et al., 2001).

#### Theory of Mind

We employed the *UCT ToM Battery* (Hoogenhout & Malcolm-Smith, 2014) as a direct measure of ToM. This battery is developmentally sequenced and includes a variety of tasks that are widely-used and well-validated (Baron-Cohen et al., 1985; Happé, 1995; Steele et al., 2003; Wellman et al., 2001). The *UCT ToM Battery* is adapted for the South African context and it has been found to be relevant to this population group (Hoogenhout & Malcolm-Smith, 2014).

The *UCT ToM Battery* tests ToM across 4 modules and 14 tasks of progressively increasing difficulty. With the exception of one task in the advanced module, all tasks use dolls and pictures to minimize demands on memory and language. The starting module was determined according to what is designated as age-appropriate. If participants passed the starting module, credit for all tasks prior to the start point was received. They continued until they failed a module, at which point the test was discontinued. A final raw score out of 100 (percentage) was calculated for each child based on all 4 modules (Ozonoff et al., 1991), even those subsequent to discontinuation. The raw scores were converted to Z-scores based on age (see Data Analysis section).

### Cognitive ability

#### Working memory

We assessed Working Memory (WM) using the backwards component of the *Numbers* subtest from the *Children’s Memory Scale* (*CMS*; Cohen, 1997). The scale score representing WM was important given the role that this cognitive ability plays in the acquisition and expression of ToM (Gordon & Olson, 1998; Ozonoff et al., 1991; Slade & Ruffman, 2005). The *CMS* has been used for South African research (Schrieff-Elson et al., 2015).

#### Intellectual functioning

General intellectual functioning (full scale IQ) was assessed as this aspect of cognition has consistently been associated with ToM (Baker et al., 2014; Happé, 1995; Hoogenhout & Malcolm-Smith, 2014; Steele et al., 2003). Verbal intellectual functioning (VIQ) was of particular interest considering that language is fundamentally intertwined with ToM development, and due to evidence of a stronger relationship between ToM and VIQ than any other aspect of general intelligence (Hamilton et al., 2016; Happé, 1995; Joseph & Tanaka, 2003). All children were assessed using the *Wechsler Abbreviated Scale of Intelligence* (*WASI*; Wechsler, 1999) given that the minimum sample age was 6 years old. The *WASI* has been translated and validated for non-Western populations (Ferret, 2011).

### Moral decision-making

We used a direct measure to assess moral decision-making in all children. The Distributive Justice task was developed at The Child NeuroSuite at the University of Chicago (Cowell et al., 2017; Huppert et al., 2019).

#### Distributive Justice task

This third-party resource allocation task assesses the subjective concept of “fairness” in moral decision-making (Huppert et al., 2019). The score is based on how children allocate resources according to their subjective perception of who is most “deserving” for conditions represented by social inequality based on wealth, merit and health. The 3 conditions were presented as 3 sets of paired images, each depicting 2 hypothetical recipients. The hypothetical recipients were represented by culturally-neutral stick-figures of the same gender as the sample in order to avoid the influence of potential social and demographic preferences. The first pair of images depicted a rich boy and a poor boy; the second pair depicted a hard-working boy and a lazy boy; and the third pair depicted a healthy and an injured boy. In each scenario, the participants were given 4 chocolates and instructed to allocate them to the hypothetical recipients according to their own judgement. The scores ranged from 0 - 4 for each condition based on the number of chocolates allocated to the morally deserving recipient (poor, hard-working, injured).

### Data Analysis

The preliminary stages of data analyses involved identifying sample characteristics and the spread of the data. Subsequently, we conducted one-tailed (directional) independent samples t-tests in order to confirm expected group differences in ToM, VIQ (represented by average scale score for verbal subtests on the *WASI*), and WM (represented by backwards component of *CMS Numbers*). Given the role that Working Memory (WM) and Verbal IQ (VIQ) play in ToM (Hamilton et al., 2016), the likely correlation between these two cognitive abilities (Conway et al., 2002) was corroborated and a composite score was created using the scale scores for the verbal subtests of the *WASI* and the scale score for Numbers from the *CMS*. VIQ and WM were equally weighted.

The raw scores (%) for ToM represented the participants’ overall achievement on the entire battery, including modules subsequent to discontinuation. To account for the effect of age on ToM development, and considering the lack of norms for the *UCT ToM Battery*, we calculated standardized *Z*-scores for theoretically informed age bands (6-7, 8-10, 11-12). Prior to this standardization of ToM scores, we checked its validity by examining the neurotypical children’s ToM performance within each age-band by descriptively comparing the neurotypical mean percentages (raw scores) with the age-appropriate expectations indicated by the literature. This was particularly important given the small sample size of each neurotypical age band. The ToM descriptive statistics for both groups are presented in Table 1 by age band.

**Table 1.** ToM Performance (Raw Scores) across Age Bands.

| Age Bands | Neurotypical |  | ASD |  |
| --- | --- | --- | --- | --- |
|  | *n* | ToM (%) | *n* | ToM (%) |
|  |  | *M(SD)* |  | *M(SD)* |
| 6 – 7 | 7 | 66.16 (13.76) | 11 | 39.56 (16.91) |
| 8 – 10 | 23 | 79.35 (7.86) | 19 | 60.71 (25.09) |
| 11 – 12 | 8 | 86.13 (4.30) | 8 | 78.76 (9.81) |

*Notes.* ASD=Autism Spectrum Disorder. ToM=Theory of Mind. *M*=Mean. *SD*=Standard Deviation.

As the ToM battery progresses from module 1 to 4 (early, basic, intermediate and advanced), the maximum score increases in increments of 25%. The mean score for neurotypical children between the ages of 6 and 7 (*M*=66.16, *SD*=13.45) indicated that children in this age group were able to understand false beliefs and some aspects of the intermediate module (possible max=75%): in line with other South African research (Robberts, 2011), these children scored higher on the strange stories (non-literal speech) than 2^nd^-order false beliefs. Participants between the ages of 8 and 10 scored a mean (*M*=79.35, *SD*=7.86) that suggested they could understand second-order false beliefs and non-literal speech in the intermediate module. Additionally, they could distinguish between lies and jokes from the advanced module to an extent, allowing them to surpass the intermediate module’s maximum score. The mean score for neurotypical children aged 11-12 (*M*=86.13, *SD*=4.3) indicated that they were competent in understanding components of all 4 modules including social faux pas (possible max=100%).

These findings are consistent with other South African literature (Robberts, 2011). Furthermore, with the exception of proficiency in strange stories prior to second-order false belief tasks, the developmental trajectory of ToM in this neurotypical sample is roughly consistent with international literature (Ackerman, 1981; Brüne & Brüne-Cohrs, 2006; Pollio & Pollio, 1979). Therefore, the neurotypical sample did not deviate from what is expected of the general population, and it was deemed appropriate to use their scores as “norms” for standardization. ASD children’s Z-scores were calculated based on the neurotypical mean scores for each age band to allow for the valid representation of ASD performance relative to age-determined neurotypical expectations. All significance testing for ToM (group differences and regression analyses) was conducted on the calculated *Z*-scores although percentages are used in reporting of results for easier interpretation.

To test for group differences in moral decision-making (hypothesis 1), we set up a mixed-design ANOVA (2X3 design; 2 groups (between-groups factor) and 3 distributive conditions (within-group factor)). Due to our clear predictions, we conducted planned comparisons (Rosenthal & Rosnow, 2008) using the raw scores indicating the number of chocolates allocated to “morally deserving” recipients. Given that the data was normally distributed, the group sizes were equal, and no assumptions were violated, the ANOVA analysis was deemed appropriate.

We also conducted some exploratory investigation of the patterns of resource allocation to gain insight into potential group differences in equitable and equal allocation strategies. To test the equity hypothesis, we determined what proportion of children from each group allocated more than half (equitable resource allocation) to the morally deserving recipient (>2) or allocated half or less to the morally deserving recipient (≤2). In order to determine whether resource allocation was contingent on group for equitable resource allocation strategies, we conducted a chi-squared test of contingency on this categorical data. Similarly, in order to explore group differences in equal allocation strategies, we investigated what proportion of children from each group shared exactly 2 chocolates to each recipient. A second chi-squared contingency analysis was run on the equal allocation frequency dataset to determine whether the decision to allocate equally was contingent on group across the distributive conditions of wealth, merit and health. More specifically, this 2 (group) by 3 (distributive conditions) design allowed us to test whether children with ASD were significantly more likely than neurotypical children to share equally across conditions. Given the small sample size of this extracted dataset, the maximal likelihood ratio was used to assess goodness of fit and to select the best model (Field, 2009). These two exploratory significance tests and accompanying descriptive statistics were used to compare equitable versus equal sharing strategies across groups, and to examine the proposed role of systemizing in ASD.

Stage 2 of the main analyses involved testing Hypothesis 2 by identifying whether ToM was a predictor of moral decision-making using Multiple Regression Analysis (MRA). A composite score for moral decision-making was created to represent the mean distributive allocation for all three distributive conditions (wealth, merit, health). One-tailed significance tests were used to examine zero-order correlations given the directional study hypotheses. In addition to ToM, variables hypothesized to impact moral decision-making were included as potential predictors in the model if they were also significantly correlated with the outcome in our sample. Alpha was set to convention (α = .05) for all significance tests.

## Results

### Sample Characteristics

Sample demographic characteristics are presented in Table 2. There were no significant group differences in demographics, namely age and SES (TFI). All participants were male and fluent in English. Therefore, the ASD and neurotypical participant groups were aggregate matched.

**Table 2.** Demographic Sample Characteristics and Descriptive Statistics across Groups.

| Characteristic | Descriptive<br>Statistic | Group |  | Significance<br>Across Groups |  | Effect<br>size |
| --- | --- | --- | --- | --- | --- | --- |
|  |  | Neurotypical<br>(*n*=38) | ASD<br>(*n*=38) | *t* | *p* | *d* |
| Age | Age Range<br>(Years:<br>Months) | 6:07 – 12:08 | 6:01 – 12:03 | - | - | - |
|  | Age (Years)<br>*M*(*SD*) | 9.39 (1.65) | 9.31 (1.83) | 0.19 | .853 | 0.04 |
| TFI (Rands per<br>Year) | *M*(*SD*) | 408370.89<br>(157439.96) | 346418.53<br>(164423.38) | 1.68 | .098 | 0.38 |
| WM (*SS*) | Range | 5-15 | 2-14 | - | - | - |
|  | *M*(*SD*) | 10.58 (2.50) | 6.55 (3.16) | 6.16 | <.001 | 1.41 |
| VIQ (*SS*) | Range | 3.50 – 16.50 | 1 – 13.50 | - | - | - |
|  | *M*(*SD*) | 11.62 (2.53) | 7.38 (2.80) | 6.94 | <.001 | 1.59 |
| ToM (%)<sup>a</sup> | Range | 52.08 - 94.06 | 10 - 91.04 | - | - | - |
|  | *M*(*SD*) | 78.34 (10.62) | 58.39 (24.49) | 4.78 | <.001<sup>b</sup> | 1.1 |
| DJ<sup>c</sup> | Wealth<br>Condition | 3.16 (0.75) | 2.53 (0.89) | - | - | - |
|  | Merit<br>Condition | 3.53 (0.60) | 2.68 (1.16) | - | - | - |
|  | Health<br>Condition | 2.89 (0.80) | 2.53 (1.06) | - | - | - |

*Notes.* ASD = Autism Spectrum Disorder. TFI = Total Family Income (annual).
WM=Working Memory. VIQ = Verbal Intelligence Quotient. ToM. DJ = Distributive Justice
*M*=Mean. *SD*=Standard deviation.
<sup>a</sup>Significance testing was conducted on the Z-values although ToM percentages are presented here for interpretation of the means.
<sup>b</sup>Equal variances not assumed (Levene's test for equality of variances, $p < .001$ )
<sup>c</sup>Significance testing results for the Distributive Justice Task are not included in this table as an ANOVA was conducted in preference to t-tests. These results are described below.

Descriptive statistics for Working Memory (WM), Verbal IQ (VIQ) and Theory of Mind (ToM) are presented in Table 2. As described earlier, the neurotypical group met all age-appropriate developmental expectations with regard to ToM performance. Additionally, results indicated that, as expected, the neurotypical group obtained significantly higher WM, VIQ and ToM scores than the ASD group.

WM and VIQ were significantly and positively correlated (*r*=.571, *p*<.001). Therefore, the use of a composite score to represent VIQ and WM as equally weighted scaled variables was appropriate. The composite for VIQ and working memory was also significantly positively correlated with ToM (*r*=.65, *p*<.001; see Table 5), as expected.

The descriptive statistics for the Distributive Justice task are also presented in Table 2. This task represents fairness in moral decision-making across three conditions: wealth, merit, and health. The data on group differences in the allocation of chocolates across distributive conditions is represented visually in Figure 1.

**Figure 1.**
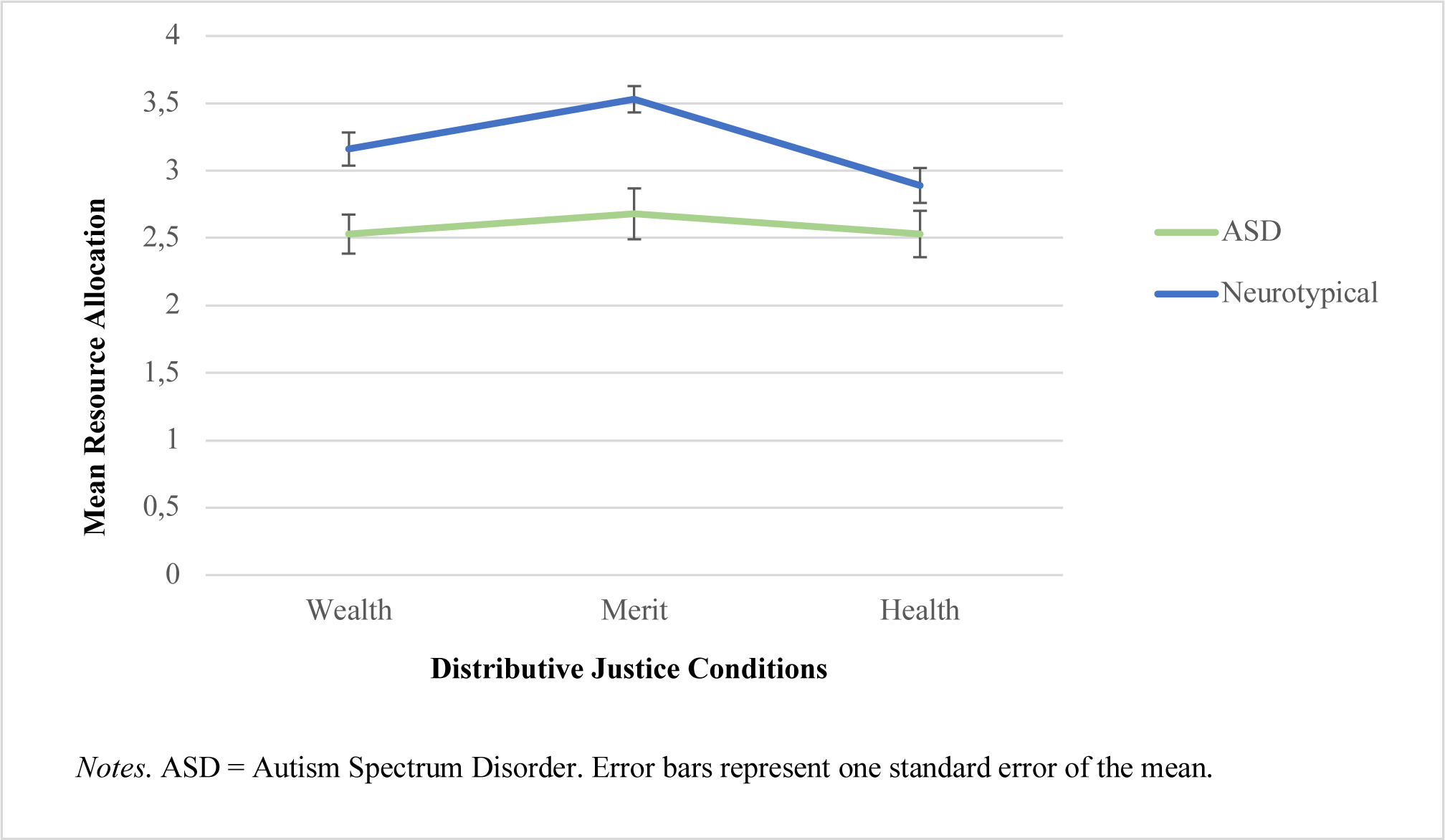
Resource allocation to the morally deserving recipient according to conditions of wealth, merit and health on the Distributive Justice task.

### Main Analyses: Stage 1

#### Group differences: Distributive Justice task

For the Distributive Justice task, we had a specific, directional hypothesis (Hypothesis 1) whereby we expected neurotypical children to use the social information and, therefore, allocate significantly more chocolates (hypothesized equitable allocation) than ASD children (hypothesized equal allocation) to the “morally deserving” recipient across conditions represented by inequality. Therefore, we conducted planned comparisons using a mixed-ANOVA design (Field, 2009). The 2 groups comprised of ASD and neurotypical children, respectively. The within-group factor was represented by the 3 distributive conditions: wealth, merit and health.

To identify whether neurotypical children allocated significantly more chocolates to the morally deserving recipient, we conducted a simple planned comparison for Group (between-subjects factor). However, we conducted a polynomial planned comparison for the Distributive Justice conditions (within-subject factor) rather than a simple planned comparison. This is because, although we did not necessarily expect there to be any differences across conditions (within-subject factor), the trend analysis of the plotted data (see Figure 1), indicated a quadratic relationship (Field, 2009). Therefore, the analysis was run with the expectation of a quadratic effect.

The planned comparison results indicated that there was no significant linear or quadratic interaction between the groups and distributive conditions. Therefore, the main effects for the contrasts were interpreted in isolation. There was a significant quadratic main effect for the distributive conditions, *F*(1, 74)=8.51, *p*=.005. Based on the priori nature of the planned comparisons, the results indicated that all the participants allocated significantly more chocolates to the morally deserving option on the merit condition than the wealth and health conditions. The effect size indicated a quadratic relationship explains 10.3% (*Partial Eta Squared*=.103) of the variance in performance on the Distributive Justice task.

The Group contrast indicated a main effect whereby neurotypical participants allocated significantly more chocolates to the morally deserving recipient than the ASD participants across conditions of wealth, merit and health, *F*(1, 74)=17.89, *p*<.001. The effect size indicated that group explains 19.5% (*Partial Eta Squared* η^2^_p_ = .195) of the variance in performance on the Distributive Justice task.

#### Patterns of resource allocation: Distributive Justice task

Upon establishing a group difference in resource allocation, we conducted further exploratory analyses to gain more insight into the pattern of allocation between groups. In order to explore group differences in equitable allocation strategies, we created distributive categories to represent a dichotomous allocation whereby the participants’ performance was either characterised as allocating more than half to the morally deserving recipient (>2) or allocating half or less to the morally deserving recipient (≤2). We calculated the proportion (%) of participants who allocated according to these categories for each group across the conditions of wealth, merit and health (see Table 3).

**Table 3.**
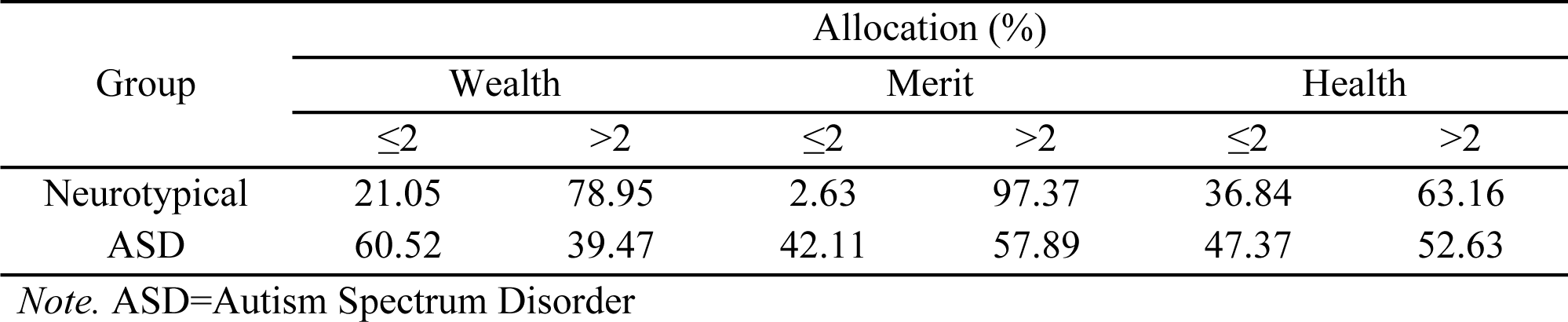
Equitable Allocation (%) across Distributive Conditions.

| Group | Allocation (%) |  |  |  |  |  |
| --- | --- | --- | --- | --- | --- | --- |
|  | Wealth |  | Merit |  | Health |  |
|  | ≤2 | >2 | ≤2 | >2 | ≤2 | >2 |
| Neurotypical | 21.05 | 78.95 | 2.63 | 97.37 | 36.84 | 63.16 |
| ASD | 60.52 | 39.47 | 42.11 | 57.89 | 47.37 | 52.63 |

*Note.* ASD=Autism Spectrum Disorder

Across all conditions, more than 60% of the neurotypical children allocated more than half (>2) of the chocolates to the morally deserving recipient. This was particularly evident on the merit condition (97.37%). In comparison to the neurotypical group, a smaller proportion of ASD children allocated more than half (>2) of the chocolates to the morally deserving recipient across all conditions, and a much larger proportion of ASD children allocated equally or gave less than half for the wealth and merit condition.

Based on our priori prediction (hypothesis 1) and the supporting descriptive pattern of resource allocation, we expected resource allocation to be dependent on group with neurotypical children being more likely to allocate equitably (i.e. more than half of the chocolates) to the morally deserving recipient. Therefore, one-sided significance testing was conducted. The results indicated that the decision to allocate more than half to the morally deserving recipient (>2) or not (≤ 2) was contingent on group for the wealth (χ^χ^(1,200) = 33.07, *p* < .001) and merit conditions (χ^χ^(1,200) = 43.61, *p* < .001). Therefore, neurotypical participants were significantly more likely to allocate more chocolates to the morally deserving recipient while ASD participants were significantly more likely to allocate half (equally) or less. The effect size for this relationship was moderate for both wealth and merit (*Cramer’s V*=.41 and .47, respectively; (Field, 2009)). However, the decision to allocate equitably (i.e. more than half) to the morally deserving recipient or not was not dependent on group for the health condition, χ^χ^(1,200) = 2.05, *p* = .099. The effect size was small (*Cramer’s V*=.10). These results are represented in Figure 2.

**Figure 2.**
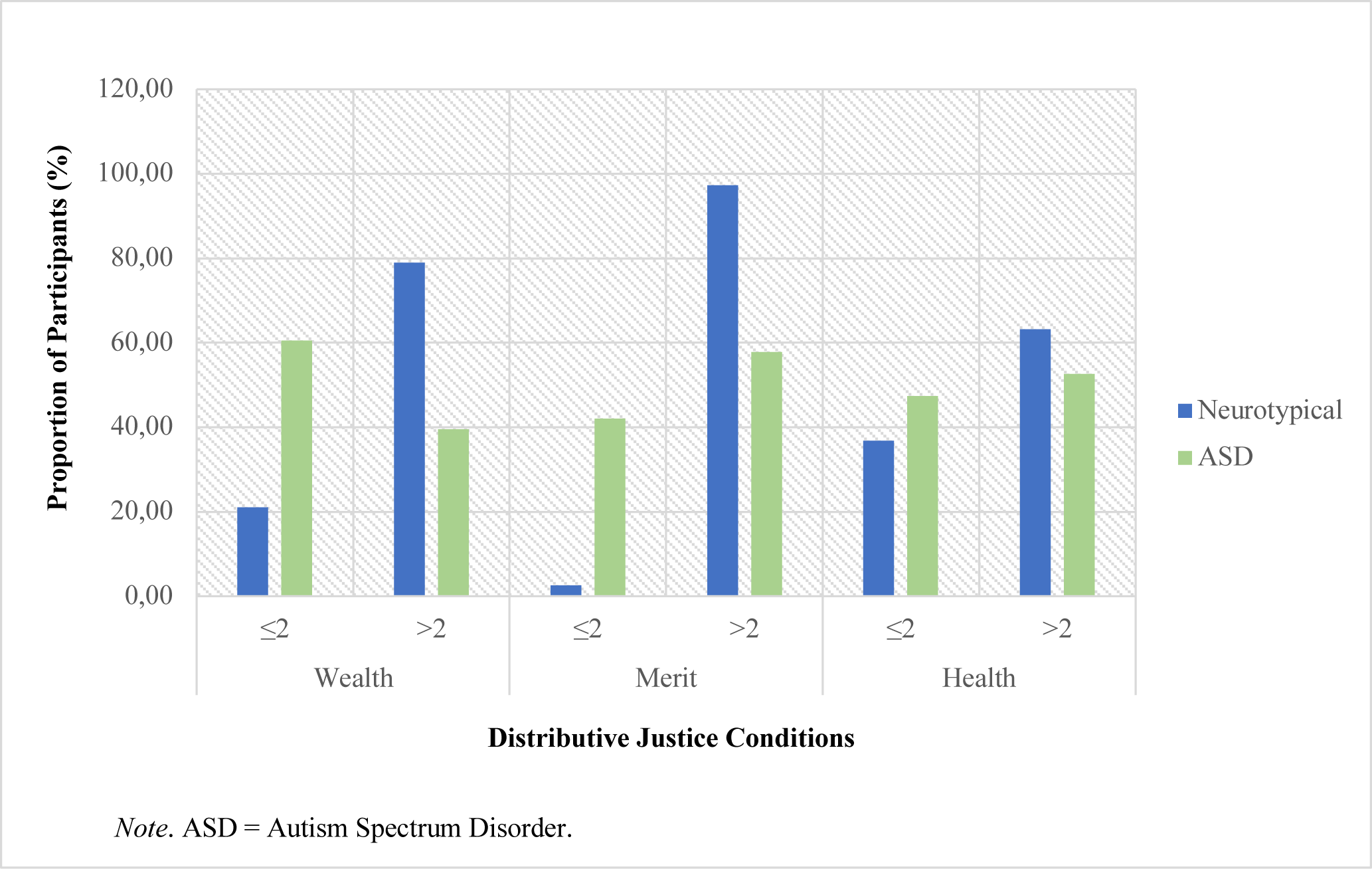
Equitable distributive resource allocation across conditions on the Distributive Justice task.

To explore group differences in equality-based resource allocation strategies, we determined what proportion of neurotypical and ASD children shared equally (See Table 4). The descriptive statistics revealed that a comparatively larger proportion of ASD children demonstrated a preference for equality. For example, on the wealth and merit conditions, a much larger proportion of ASD children (57.89%, 31.58%) allocated resources equally in comparison to neurotypical children (21.05%, 2.63%).

**Table 4.** Equal Allocation (%) Across Distributive Conditions.

| Group | Equal Allocation (%) |  |  |
| --- | --- | --- | --- |
|  | Wealth | Merit | Health |
| ASD | 57.89 | 31.58 | 34.21 |
| Neurotypical | 21.05 | 2.63 | 36.84 |

Note. ASD=Autism Spectrum Disorder

While the initial chi-squared analysis (see Figure 2) corroborated that neurotypical children are significantly more likely to allocate resources equitably, there was also an overall trend (see Figure 1) towards equitable allocation in the ASD group. However, this was significantly weaker than the neurotypical group due to a large proportion of children with ASD allocating equally (see Table 4). Given the a priori expectation that some preference for equality may be evident in the ASD group (component of hypothesis 1) and based on the supporting descriptive pattern of resource allocation, a second chi-squared analysis test was conducted on the equal allocation data. The objective of this was to determine whether equal resource allocation was contingent on group across distributive conditions. Frequency data on equal allocation decisions was extracted to allow for a specific focus on differences in equal sharing decisions across groups. The results of chi squared contingency analysis (maximum likelihood ratio) indicated that children with ASD were significantly more likely to allocate equally than neurotypical children across distributive conditions of wealth, merit and health, χ^χ^(2, 185) = 24.31, *p* < .001. The effect size for this relationship was moderate (*Cramer’s V*=.35; (Field, 2009). These results are represented in Figure 3.

**Figure 3.**
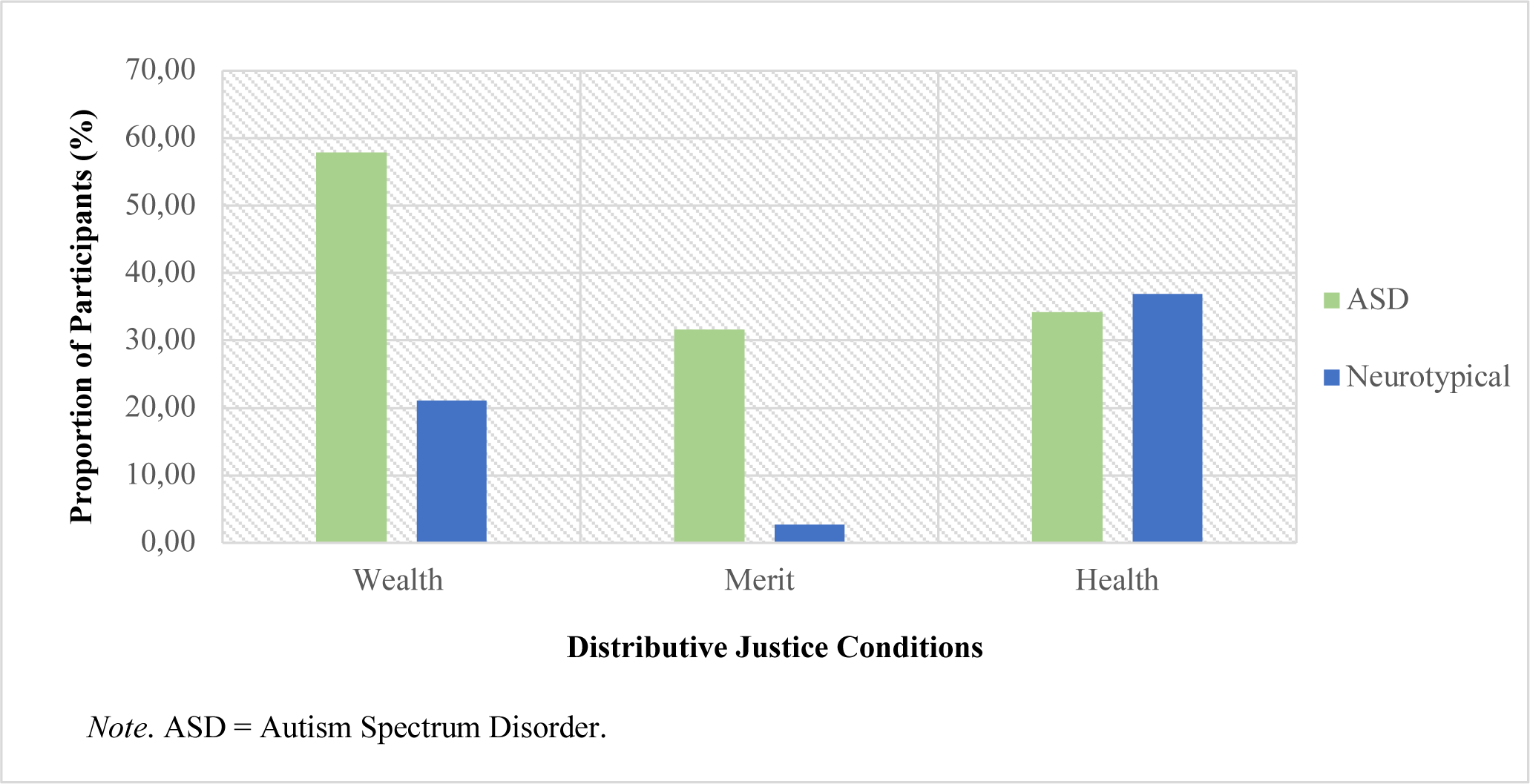
Equal distributive resource allocation across conditions on the Distributive Justice task.

Therefore, while the majority of neurotypical children allocated equitably, the decision to allocate equally was significantly more frequent with a comparatively large proportion of the ASD group adopting this strategy. The discrepancy between groups was less apparent on the health condition where a similar proportion of children allocated equally in both the neurotypical (36.84%) and ASD (34.21%) groups. This is due to a change in the neurotypical children’s response pattern with a shift from allocating more than half to allocating equally.

### Main Analyses: Stage 2

For the second stage of data analysis, we aimed to explore whether differing group characteristics predicted moral decision-making. More specifically, we built hierarchical MRA models to identify whether ToM was a predictor of moral decision-making as proposed by Hypothesis 2. For MRA, moral decision-making was represented by a composite score created using the mean of all three distributive justice conditions (wealth, merit, and health). All zero-order correlations of interest are represented in Table 5.

**Table 5.** Zero-Order Correlation Matrix.

|  | 1. Age<br>(Months) | 2. SES<br>(TFI) | 3. WM/VIQ<br>(Composite SS) | 4. ToM<br>(Z) | 5. Interaction<br>(ToM*VIQ/WM) | 6. DJ<br>(Composite) |
| --- | --- | --- | --- | --- | --- | --- |
| 1 | - |  |  |  |  |  |
| 2 | -0.04 | - |  |  |  |  |
| 3 | 0.12 | 0.10 | - |  |  |  |
| 4 | 0.05 | 0.22* | 0.65*** | - |  |  |
| 5 | 0.34*** | 0.23* | 0.80*** | 0.85*** | - |  |
| 6 | 0.17 | 0.26* | 0.30** | 0.37*** | 0.43*** | - |

*Notes.* SES=Socio-Economic Status. TFI=Total Family Income (annual). WM=Working Memory. VIQ=Verbal Intelligence Quotient. ToM=Theory of Mind. DJ=Distributive Justice. Z=Z scores. SS=Scale Scores
\*. Correlation is significant at the .05 level (1-tailed).
\*\*. Correlation is significant at the .01 level (1-tailed).
\*\*\*. Correlation is significant at the .001 level (1-tailed)

### MRA model building

Initially, age, SES, VIQ/WM and the interaction between VIQ/WM and ToM were considered as potential predictors of moral decision-making. Given that group was expected to be strongly correlated with almost every predictor of interest, it was not included in the MRA model to preliminarily limit multicollinearity. Instead, ToM was considered according to a spectrum with neurotypical children scoring on the high end and ASD children scoring on the low end (see Table 1 and 2).

Based on the significant zero-order correlation between ToM and the outcome variable (moral decision-making; *r*=.37, *p*<.001), this relationship was worth further investigation. In building the model, ToM was entered subsequent to other potential predictors whose theoretical hypothesized association with moral decision-making was corroborated by a significant correlation. The purpose of this hierarchical MRA model was to determine whether ToM had a predictive effect over and above the effect of other potential predictors.

Age was not significantly correlated with the composite score for moral decision-making (*r*=.17, *p*=0.07). Therefore, it was excluded from the models. However, SES was significantly correlated with moral decision-making (*r*=.26, *p*=0.01) and was recognized as a potential predictor for consideration in the model. Similarly, the interaction between VIQ/WM and ToM was significantly correlated with moral decision-making (*r*=.43, *p*<.001) and it was initially considered in the model and entered subsequent to VIQ/WM and ToM, respectively. However, considering the significant inter-correlation (*r*=.65, *p*<.001) between VIQ/WM and ToM and the known relationship between these variables (Hamilton et al., 2016), emphasis was placed on assessing multicollinearity (see supplementary material). This assessment indicated that inclusion of VIQ/WM was creating a multicollinearity concern due to the large amount of shared variance between VIQ/WM, ToM, and the interaction between these two variables. All models built subsequent to the removal of VIQ/WM and the interaction between VIQ/WM and ToM, did not present a problem of multicollinearity.

Based on the zero-order correlations and hypothesized effects, SES was entered followed by ToM. This model was significant, *F*(2, 73) = 7.48, *p*=.001. The adjusted effect size indicated that overall, this model (see Table 6) explained 14.7% of the variance (*R*^2^= .170, *adj*. *R*^2^= .147). The inclusion of ToM made a significant contribution by explaining an additional 10.4% (*R*^2^*c*ℎ*ange* =.104) of the variance in moral decision-making, *F*(1,73)=9.13, *p*=.003. ToM also emerged as a significant predictor (*β*=.33, *t*=3.02, *p*=0.003). Although SES (*β*=.19, t=1.7, *p*=0.093) was not an individually significant predictor, it was included in the model as it still explains a small, yet significant proportion (5.4%) of the variance (*R*^2^*c*ℎ*ange* = .066, *adj R*^2^*c*ℎ*ange* = .054)

**Table 6.** Predictors of Moral Decision-Making on the Distributive Justice Task.

| Model | *R* | *R Square* | *Adjusted R Square* | Std. Error of the Estimate | Change Statistics |  |
| --- | --- | --- | --- | --- | --- | --- |
|  |  |  |  |  | *R Square Change* | *Sig. F Change* |
| 1 | .257 | .066 | .054 | .68 | .066 | .025 |
| 2 | .412 | .170 | .147 | .65 | .104 | .003 |

*Notes.*
Model 1: Constant, Socio-Economic Status (SES; Total Family Income (TFI))
Model 2: Constant, SES (TFI), Theory of Mind (ToM; Z)

## Discussion

The results of our study provide evidence for significant group differences in moral decision-making between neurotypical children and children with ASD. Furthermore, they support the hypothesized predictive effect of ToM on moral decision-making.

### Differences in Moral Decision-Making Between Groups

The Distributive Justice task (a measure of moral decision-making regarding the subjective perception of fairness) results revealed a significant between-groups difference (ANOVA planned comparison; main effect for group) whereby the neurotypical children allocated significantly more resources to the morally deserving recipient across conditions representing social inequality based on wealth, merit and health. The significant group difference was further supported by a clear distinction in ASD and neurotypical children’s pattern of resource allocation. Although both groups demonstrated an overall tendency to allocate equitably (both groups had a mean sharing score of more than half across conditions), this trend was significantly stronger in the neurotypical group; exploratory analyses revealed that the decision to allocate more than half of the resources to the morally deserving recipient or not, was contingent on group for the wealth and merit conditions. Therefore, neurotypical participants were significantly more likely than children with ASD to allocate more resources to the morally deserving recipient (equity-based sharing), particularly when faced with scenarios of inequality based on wealth or merit.

In exploring the patterns of resource allocation further, we found that while the majority (more than 60%) of the neurotypical children allocated more than half of their resources (equitable distribution) to the morally deserving recipient across conditions, a comparatively large proportion of the children with ASD demonstrated a preference for equality. Furthermore, it was found that the decision to allocate exactly half of the resources was contingent on group whereby children with ASD were significantly more likely to share equally than neurotypical children across conditions.

Although it is apparent that both groups demonstrated an overall preference for equity, there is a group difference characterised by a significantly increased likelihood of neurotypical children allocating equitably, particularly on the wealth and merit conditions. Additionally, despite the overall trend in the direction of equity in the ASD group, a comparatively larger proportion of children with ASD chose to share equally in comparison to neurotypical children. Therefore, the decision to allocate equally was significantly more frequent among children with ASD. Our results suggest that neurotypical children mostly interpret fairness within the context of social information to make equitable distributions while children with ASD allocate significantly less to the morally deserving recipient due to a comparatively increased tendency to equate fairness with equality.

The neurotypical children’s performance in our study is consistent with previous research demonstrating that, by the age of 6, children typically start considering social norms and making equitable decisions about fairness (Damon, 1977; Rizzo et al., 2016; Rizzo & Killen, 2016). This was particularly evident on the needs-based (wealth) and performance-based (merit) resource allocation items. For example, the neurotypical children’s preference for equity on the wealth condition is in alignment with previous studies indicating that children share more with people in need (Malti et al., 2016; Paulus, 2014) and give more to recipients of low wealth characterised by smaller houses (Shutts et al., 2016). The value that neurotypical children placed on hard-work and merit replicates research indicating that children share more with others who have contributed more effort to a joint task (Kienbaum & Wilkening, 2009). Although the neurotypical children’s preference for equity on the health condition was less marked (equitable resource allocation was not contingent on group), they still allocated significantly more resources to the morally deserving recipient than the ASD group. However, while the health condition is designed to evoke an empathic response based on sympathy which should become enhanced with age in neurotypical development (Malti et al., 2012; Malti et al., 2016; Paulus, 2014), it had the weakest effects in our study. Therefore, further research should reconsider this item’s validity.

In considering the mean age of the sample, the moral decision-making behaviour of a large portion of the ASD group was atypical (Rizzo & Killen, 2016). While there was an overall resource allocation trend towards equity, this was weak in comparison to the neurotypical group. Additionally, when examining the proportion of children in each group who chose to share equally, it became evident that this choice was significantly more frequent in children with ASD. This is not age-appropriate as it is consistent with what is expected from young neurotypical children who have not yet developed the ability to integrate social norms into their decision-making (Rizzo et al., 2016).

Our finding of atypical moral decision-making in a significant portion of the ASD group supports hypothesis 1, and is consistent with previous ASD literature (Gleichgerrcht et al., 2013; Li et al., 2019; Moran et al., 2011; Rogé & Mullet, 2011; Salvano-Pardieu et al., 2016; Schneider et al., 2013). Given the overall trend towards equity in the ASD group, it is possible that the subgroup of equal allocators were developmentally delayed in terms of their ability (ToM) to use and integrate social information in moral decision-making. This may have contributed toward an inflexible approach to moral decision-making, preventing such children from adapting the practical application of fairness as a moral principle. It is speculated that the reliance on rules and preference for equality in some children with ASD may be indicative of social functioning equivalent to a younger neurotypical child that may improve slowly with age (Baumard et al., 2012; Smith et al., 2013).

However, based on the E-S Theory (Baron-Cohen, 2009; Baron-Cohen et al., 2003) and the EMB Theory of Autism (Baron-Cohen, 2002), it is also proposed that children with ASD who selected to distribute resources equally may be systemizing rather than empathizing. In this manner, ASD children could have considered each social scenario presented to them as a fixed “system”. Using this approach, it is possible that they used rules to respond to scenarios that they perceived to be lawful and predictable. Given the limited predictive power of systemizing relative to empathizing in the social world (Baron-Cohen, 2009; Baron-Cohen et al., 2003), this type of approach in ASD could potentially result in moral decision-making behaviour that society deems inappropriate (Dunfield et al., 2013; Shaw et al., 2012). In theory, this increased reliance on uncompromising rules that are not easily adjusted to contextual social information is considered to be a core defining feature of social functioning in ASD rather than an indicator of developmental delay. Longitudinal research is necessary to tease out the developmental trajectory of ToM and its role in moral decision-making within the context of ASD.

An additional finding that emerged from the Distributive Justice task was the differing performance across conditions of wealth, merit and health. Although neurotypical participants showed a preference for equity over equality based on performance (merit) and need (wealth and health), the strength of this relationship varied across conditions.

Firstly, the decision to allocate equitably was not contingent on group for health as there was less group disparity on this condition; a similar proportion of children divided equally in both the neurotypical (36.84%) and ASD (34.21%) groups. This seems to be due to a change in the neurotypical children’s response pattern with a shift from sharing equitably to sharing equally. Therefore, it is possible that the health condition did not evoke the same sense of justice in the neurotypical children as the wealth and merit conditions did. Therefore, injury appears to be a less motivating recipient cue than material need and hard work with increased ambiguity around the identification of the morally-deserving recipient. The health condition’s limited ability to evoke empathy across groups is possibly due to the nature of the injury not being considered dire by school children who may even enjoy the attention and special privileges associated with having a cast. Additionally, due to the nature of the resources, moral decision-making on this condition may also have been influenced by practical thinking whereby chocolates are not considered to be healthy for a person who is injured or unwell.

Secondly, there was a significant quadratic effect pertaining to the distributive conditions of the Distributive Justice task whereby all children allocated significantly more resources to the morally deserving recipient on the merit condition than the wealth and health conditions. Therefore, children placed greater value on hard work (performance) than low wealth or injury (material and emotional need). Unlike the wealth and health condition, the merit condition represented a social scenario where the recipients were directly responsible for their actions. Therefore, performance on this condition may be impacted by the children’s understanding of the link between choices and consequences (Kienbaum & Wilkening, 2009; Rizzo et al., 2016). Furthermore, it appears that the degree to which they made equitable moral decisions could have been dependent on the extent of the participant’s rational reasoning as well as their sense of social responsibility (Kienbaum & Wilkening, 2009; Rizzo et al., 2016). This potentially lends support to a preference for performance-based equity over needs-based equity due to a more clear-cut perception of justice.

This specific manifestation of fairness may also reflect South African cultural norms given that unique social environments provide different opportunities for social learning which, in turn, influences the development of social cognition (Vredenburgh et al., 2017). While individualistic cultures prioritize autonomy and personal goals, collectivist cultures prioritize integrated family structures and a sense of community (Triandis, 1995, 2001). Typically, African countries are considered to be collectivist (Triandis, 1989). However, South Africa has a diverse population and this cultural affiliation may not be generalizable to all groups of people. Recent cross-cultural research indicates that South Africa is more individualistic than collectivist with a Hofstede score of 65 (Huppert et al., 2019). Additionally, this study only recruited English-speaking participants, making it even more likely to be representative of individualistic cultural viewpoints (Eaton & Louw, 2000).

This is relevant considering that differences in individualism and collectivism translate into different fairness preferences. For example, children with more group-oriented goals who identify with collectivist viewpoints may be less motivated to address inequalities due to their reliance on larger social support networks (Paulus, 2015). In contrast, individualistic societies encourage competition and allocate resources in a manner that is dependent on work input (Sigelman & Waitzman, 1991). Recent research has also found that children from individualistic cultures demonstrate a preference for equitable resource allocation in social scenarios representing inequality based on wealth and merit, while children from collectivist countries demonstrate a preference for equitable resource allocation in scenarios representing inequality based on health (Huppert et al., 2019). Children from individualistic cultures also endorse equitable distributions at an earlier age. Therefore, given South Africa’s individualistic tendency, the preference for equity (particularly on the merit and wealth conditions) is consistent with previous cross-cultural literature.

The fact that both groups allocated more to the morally deserving recipient for merit than wealth and health, also suggests that this condition may be associated with a greater degree of overlap between a preference for equity in most children (particularly neurotypical children) and a tendency to systemize in the portion of children with ASD who demonstrated atypical moral decision-making. This is most likely because the rules regarding merit and reward are typically reinforced within the classroom of mainstream and special-needs schools. Therefore, the rule used by ASD children may have been clearer on this condition and more consistent with the equitable distributive response. More specifically, the equitable response may also be deemed the “empathic response.” Therefore, the application of a taught classroom rule (systemizing) may have presented in the same way as empathizing.

In sum, both groups demonstrated a preference for performance-based equity (merit) while emotional need (health) was the least motivating recipient cue in the neurotypical group. Despite these preferences, ASD and neurotypical children still differed significantly in their moral decision-making across conditions of wealth, merit and health. While both groups demonstrated an overall preference for equity, this trend was significantly stronger in the neurotypical group. Furthermore, in comparison to the neurotypical group, the ASD group was significantly more likely to allocate resources equally due to a larger proportion of children with ASD making this decision. The identification of atypical moral decision-making in ASD children characterised by a comparative preference for equality rather than equity is novel and significant due to the likely impact on social competence and social inclusion (Dunfield et al., 2013; Shaw et al., 2012). The possible mechanisms for this include developmental delay in social ability (including ToM) and an increased tendency to systemize, both of which require further investigation in future research.

### Difference in Theory of Mind between Groups

As we expected and in accordance with previous research (Baron-Cohen, 1995; Baron-Cohen et al., 1985; Happé, 1995; Happé & Frith, 1996; Hoogenhout & Malcolm-Smith, 2014), the neurotypical children scored significantly higher in ToM than the children with ASD. For the rest of the study, ToM was understood according to a spectrum with the upper end representing neurotypical participants and the lower end representing ASD participants. The conceptualisation of ASD and neurotypical children’s abilities according to one continuous spectrum is consistent with research suggesting ASD characteristics are normally distributed within the population(Constantino, 2011; Constantino & Todd, 2003), with clinically diagnosed individuals representing the severe end (Sasson et al., 2013).

### Predictors of Moral-Decision-Making

#### Theory of Mind

In support of Hypothesis 2, ToM emerged as a predictor of moral decision-making; increasing ToM ability was predictive of equitable resource allocation on the Distributive Justice task. This finding supports neuroimaging research indicating that ToM is typically implicated in moral judgement (Young et al., 2007; Young & Dungan, 2012) and decision-making (Reniers et al., 2012).

It also potentially explains differences in the way neurotypical and ASD children make moral decisions. Both ToM and moral decision-making can be conceptualised according to a spectrum with advancing ToM ability and increasing resource allocation (to a morally deserving recipient) on the upper end. The significant differences in ToM and moral decision-making between ASD and neurotypical children indicated that they represent the upper and lower bounds, respectively. Therefore, being a neurotypical child (in contrast to an ASD child) was associated with significantly higher ToM scores and, in turn, the increased allocation of resources to a morally deserving recipient (equitable distributions).

The association between atypical moral decision-making and ToM deficits in children with ASD is in line with previous research on moral judgement and reasoning in ASD adults (Gleichgerrcht et al., 2013; Moran et al., 2011; Rogé & Mullet, 2011; Salvano-Pardieu et al., 2016; Zalla et al., 2011). While it is suggested that moral decision-making in ASD is driven by an overriding interest in non-social stimuli rather than a lack of understanding, ToM skills are still considered necessary for moral decision-making by driving an intricate mechanism that integrates affective empathy with social information (Li et al., 2019).

Based on these results, we propose a conceptual model of the relationship between empathy and morality in neurotypical moral-decision-making (see Figure 4). Given what we know about moral decision-making in general, the suggested model proposes that ToM is required to integrate empathic arousal (affective empathy) with information about the mental states of others and, the consequences of behaviour within a moral context. In neurotypical children, this is likely to promote empathic concern which plays an influential role in moral decision-making. In considering how this model may be applied to the ASD population, it is worth considering research which has shown that affective empathy remains intact while cognitive empathy is impaired (Dziobek et al., 2008; Fan et al., 2014). Therefore, in line with the results of this study, it is likely that the evident ToM (cognitive empathy) deficits or delays in the ASD group may be disrupting the typical process of moral decision-making. More specifically, we propose that ToM deficits may have impaired perspective-taking and prevented the effective integration of empathic arousal and emotional sharing (affective empathy) with social information in a moral context. As a result, the children with ASD may have had difficulty translating their empathic arousal into empathic concern and, in turn, may have been less motivated to behave “empathically” in making moral decisions.

**Figure 4.**
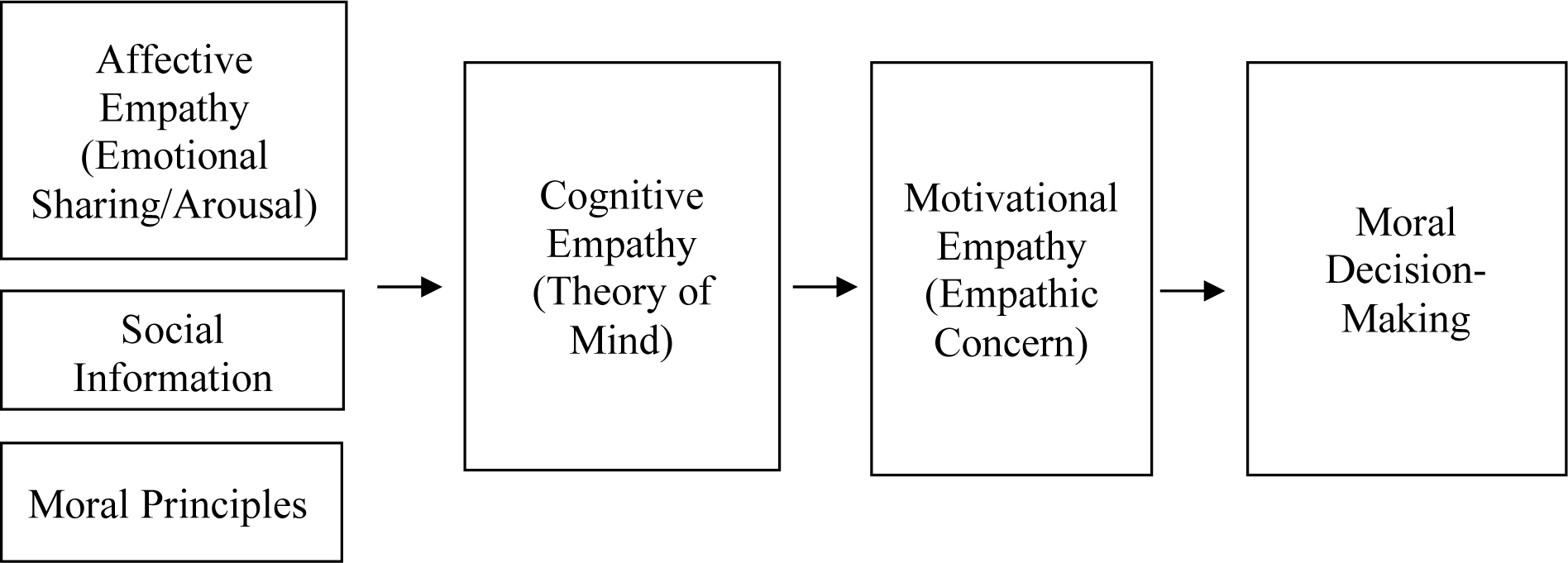
A proposed model of the relationship between empathy and moral decision-making.

In line with the E-S Theory, it is possible that some children with ASD may have consequently resorted to systemizing and applied concrete rules that they had either learned or self-developed to the social situation. Therefore, we do not propose a lack of morality in ASD but rather a difficulty using social information in the application of moral principles due to ToM deficits. The relevance of this finding lies in its ability to aid people’s understanding of behaviour in ASD which is often misinterpreted and stigmatised in the social context.

The support that our study provides for the establishment of the proposed model (see Figure 4) is relevant considering the sequential and functional nature of the model and its potential ability to make predictions about moral decision-making based on empathy deficits. While our study has provided quantitative evidence for atypical moral decision-making in ASD, the role of systemizing vs. empathizing is less clear. However, the possible link between ToM deficits and systemizing in ASD provides a platform for further research on this topic. This type of research could not only aid people’s understanding of social impairment in ASD, but also provide guidance for intervention strategies that recognise ToM deficits in ASD and acknowledge the use of therapeutic techniques that use systemizing (fixed input-operation-output relationships) as a tool to improve social skills.

### Limitations and Considerations for Future Research

#### Sample

The sample size and demographic variability of this study was limited to children fluent in English as the *ADOS2* has not yet been formally translated or validated for other African languages. This impacts on the extent to which results can be generalised to the broader population. Nonetheless, the study provides data from a non-western context which is needed, particularly in ASD research (de Leeuw et al., 2020). While the sample size was limited by design factors, *a priori* power analyses indicated sufficiency. Additionally, although some of the neurotypical age-groups were small, their ToM performance corresponded with both international and South African literature indicating valid standardisation for normative scores.

#### Confounding variables

While SES was considered in the study, we were limited in our ability to fully explain its influence on moral decision making. SES contributed a significant portion of the predictive model’s variance and it was positively correlated with increased equitable resource allocation. However, it did not emerge as a significant individual predictor – therefore its role requires further investigation. Additionally, for valid interpretation, future research should use multiple indicators of SES rather than TFI alone as this is considered to be more appropriate for research in developing countries such as South Africa (Cooper et al., 2012).

This study was also limited in that it did not account for additional potential confounding variables such as executive functioning, verbal skills, and IQ. Given that ToM is intricately developmentally intertwined with all of these variables, there is ongoing debate regarding the directionality of this complicated, inter-dependent relationship (Gordon & Olson, 1998; Ozonoff et al., 1991; Slade & Ruffman, 2005). Statistically, it is difficult to tease out their individual effects as they tend to be significantly inter-correlated with one another. This study faced the same challenges and, due to multicollinearity concerns, the WM/VIQ composite was omitted from the MRA model; further research is required to investigate the full nature and direction of this relationship. Lastly, given that self-regulation has also been suggested as key component of empathy in morality (Decety, 2011) and identified as a significant predictor of moral decision-making (Cowell et al., 2017; Smith et al., 2013), further insight into its role in the proposed 3-pillar model is recommended.

Longitudinal research also has the potential to provide further insight into the role of ToM in moral decision-making by determining whether such deficits remain a permanent feature of Autism Spectrum Disorder, or form part of a developmentally delayed picture.

## Summary and Conclusions

In comparison to children with ASD, neurotypical children allocated significantly more resources to the morally deserving recipient on all conditions of the Distributive Justice task. While both groups demonstrated an overall preference for equitable allocations, this trend was significantly stronger in the neurotypical group, particularly on the wealth and merit conditions. In line with individualistic cultural norms, this is indicative of a preference for equity-based distributions. In contrast, while children with ASD demonstrated an overall tendency towards equity-based sharing, they were significantly more likely than neurotypical children to allocate equally due to this decision being more frequent in a comparatively larger portion of the ASD group. ToM emerged as a significant predictor of moral decision-making with higher ToM scores being predictive of increased equity-based sharing in scenarios representing social inequality. It is speculated that ToM (cognitive empathy) deficits in ASD may underly an atypical preference for equality in moral decision-making due to limited integration of empathic arousal (affective empathy) and moral information. However, further longitudinal research is necessary to identify whether this is a permanent feature of the disorder or due to delayed ToM development. This study lends support for the establishment of a predictive model for moral decision-making on the basis of sequentially related empathy components, and provides a rationale for future research investigating the role of empathizing vs. systemizing in moral decision-making, within the context of ASD.

## Declarations

### Authors’ Contributions

Jessica Ellen Ringshaw is the primary author of this paper which was conducted in affiliation with the University of Cape Town. Data collection for the study was conducted by both Jessica Ellen Ringshaw and Katie Hamilton. Study conception and design, material preparation, data analysis, and the interpretation of results was conducted by Jessica Ellen Ringshaw with input and supervision from Susan Malcolm-Smith and Katie Hamilton. Jessica Ellen Ringshaw drafted and revised the manuscript with guidance and critical review from Susan Malcolm-Smith and Katie Hamilton. All authors provided approval of the final version to be published.

### Ethical Considerations

This study was approved by the University of Cape Town’s institutional Faculty of Health Sciences Human Research Ethics Committee (HREC reference: 346/2017) and the Ethics Committee of the Faculty of Humanities (reference: PSY2016-015). The authors of this study certify that it was conducted in accordance with the ethical standards as laid down in the 1964 Declaration of Helsinki, as revised in 2013.

## Data Availability

The data associated with this manuscript has not been deposited into a data repository and will not be available prior to peer-reviewed publication.

## Acknowledgements

We thank Professor Jean Decety and the Child NeuroSuite at the University of Chicago for making the Distributive Justice Task available for use in this study. We would also like to thank Kirsty Carter for her support throughout the research process as a fellow ASD researcher. The authors did not receive support from any organisation for the submitted work. No funding was received for conducting this study or to assist with the preparation of this manuscript. During her MA degree, while conducting this research, Jessica Ellen Ringshaw was supported by a Masters Research Scholarship from the University of Cape Town. Katie Hamilton has received support from the Harry Crossley Foundation and National Research Foundation for her research work.

