## Supplementary Material for "Theory of Mind and Moral Decision-Making in the Context of Autism Spectrum Disorder"

+27 84 4114084

4th Floor, Institute of Child Health Building, Red Cross War Memorial Children's Hospital, Klipfontein Road, Rondebosch, Cape Town, South Africa, 7700

### Supplementary Material

#### The Relationship Between VIQ, WM, and ToM in Moral Decision-Making Analyses

Verbal IQ (VIQ), Working Memory (WM) and Theory of Mind (ToM) are developmentally intertwined and statistically inter-correlated. Multicollinearity assessments were conducted to determine whether these variables (VIQ/WM composite and ToM) could be validly included as potential predictors for moral decision-making in hierarchical multiple regression analyses. While the multicollinearity assessment justified the removal of VIQ/WM from MRA analyses, we acknowledge the complicated role it plays in ToM.

***Multicollinearity.*** Multicollinearity was initially assessed based on the full proposed model for moral decision-making using all predictors (SES, VIQ/WM, ToM, VIQ/WM-ToM interaction) that correlated with the outcome variable (Distributive Justice composite; see Tables 7 -10). Collinearity statistics (see Table 7) raised concern as the average VIF values for WM/VIQ, ToM, and the interaction between VIQ/WM and ToM were considerably higher than 1 and above the cut-off of 2.5 (Senaviratna & Cooray, 2019). Of particular concern is the interaction between VIQ/WM and ToM, given that the associated VIF value is higher than the cut-off of 5 (Kim, 2019; Yu et al., 2015) and the tolerance value is below 0.2 (Menard, 1995). Collinearity diagnostics (see Table 8) revealed unstable regression models as some Eigenvalues were much larger than others, and a few (dimensions 4 and 5) raised concern for multicollinearity as the eigen values were close to zero (Kim, 2019; Senaviratna & Cooray, 2019). Similarly, the corresponding condition indices were large with those above 10 (dimension 4) and even 15 (dimension 5; Midi et al., 2010) indicating multicollinearity. Collinearity diagnostics indicate that a high proportion of variance (>50%; Belsley, 1991)) in both ToM (63%) and the interaction between ToM and VIQ/WM (95%) loaded onto the same small eigen value along dimension 5. Therefore, the interaction between ToM and VIQ/WM was removed.

Table 7.

*Collinearity Statistics for the Proposed Moral Decision-Making Model 1*

| Predictor Variables | Tolerance | VIF |
|---|---|---|
| SES (TFI) | .93 | 1.08 |
| WM/VIQ Composite | .35 | 2.86 |
| ToM (*Z*) | .28 | 3.54 |
| Interaction (VIQ/WM*ToM) | .17 | 5.85 |

*Notes.* SES=Socio-Economic Status. TFI=Total Family Income. WM=Working Memory. VIQ=Verbal Intelligence Quotient. ToM=Theory of Mind.

Table 8.

*Collinearity Diagnostics for the Proposed Moral Decision-Making Model 1*

| Dimension | Eigen Value | Condition Index | Variance Proportions | | | | |
|---|---|---|---|---|---|---|---|
| | | | (Constant) | SES (TFI) | VIQ/WM (Composite) | ToM (*Z*) | Interaction (VIQ/WM* ToM) |
| 1 | 3.88 | 1.00 | .00 | .01 | .00 | .00 | .00 |
| 2 | 0.95 | 2.02 | .00 | .00 | .00 | .22 | .00 |
| 3 | 0.13 | 5.50 | .00 | .83 | .05 | .02 | .01 |
| 4 | 0.02 | 12.87 | .53 | .16 | .69 | .13 | .04 |
| 5 | 0.02 | 16.23 | .46 | .00 | .26 | .63 | .95 |

*Notes.* SES=Socio-Economic Status. TFI=Total Family Income. WM=Working Memory. VIQ=Verbal Intelligence Quotient. ToM=Theory of Mind.

After removing the interaction between Tom and VIQ/WM, a second collinearity assessment was conducted to determine whether the multicollinearity problem had been resolved (see Table 9). While the VIF and tolerance values had improved, the collinearity diagnostics still indicated the presence of multicollinearity. The regression model was found to be unstable with some Eigenvalues being much larger than others (see Table 10). Dimension 4 raised concern for multicollinearity as it's eigen value was close to zero (Kim, 2019; Senaviratna & Cooray, 2019) and its condition index was over 10 (Midi et al., 2013). Furthermore, a high proportion of variance (>50%; Belsley, 1991) in both ToM (51%) VIQ/WM (84%) loaded onto the same small eigen value along dimension 4. Therefore, VIQ/WM was not included in any MRA analyses as there was too much shared variance with ToM to reliably assess the unique predictive ability of ToM. Removal of these predictors resolved the multicollinearity issue in all further models.

Table 9.

*Collinearity Statistics for the Proposed Moral Decision-Making Model 2*

| Predictor Variables | Tolerance | VIF |
|---|---|---|
| SES (TFI) | .95 | 1.052 |
| WM/VIQ Composite | .58 | 1.73 |
| ToM (*Z*) | .56 | 1.79 |

*Notes.* SES=Socio-Economic Status. TFI=Total Family Income. WM=Working Memory. VIQ=Verbal Intelligence Quotient. ToM=Theory of Mind.

Table 10.

*Collinearity Diagnostics for the Proposed Moral Decision-Making Model 2*

| Dimension | Eigen Value | Condition Index | Variance Proportions | | | |
|---|---|---|---|---|---|---|
| | | | (Constant) | SES (TFI) | VIQ/WM (Composite) | ToM (*Z*) |
| 1 | 3 | 1.00 | .00 | .01 | .01 | .01 |
| 2 | 0.86 | 1.87 | .00 | .00 | .01 | .47 |
| 3 | 0.12 | 5.03 | .02 | .81 | .15 | .01 |
| 4 | 0.02 | 11.44 | .98 | .17 | .84 | .51 |

*Notes.* SES=Socio-Economic Status. TFI=Total Family Income. WM=Working Memory. VIQ=Verbal Intelligence Quotient. ToM=Theory of Mind.
